# Containing pandemics through targeted testing of households

**DOI:** 10.1101/2020.10.30.20219766

**Authors:** André Voigt, Nikolay Martyushenko, Emil Karlsen, Martina Hall, Kristen Nyhamar, Stig William Omholt, Eivind Almaas

## Abstract

While invasive social distancing measures have proven efficient to control the spread of pandemics in the absence of a vaccine, they carry vast societal costs. Guided by the finding that large households function as hubs for the propagation of COVID-19, we developed a data-driven individual-based epidemiological network-model to assess the intervention efficiency of targeted testing of larger households. For an outbreak with reproductive number *R* = 1.5, we find that weekly testing of just the 15% largest households is capable of forcing *R* below unity. For the case of *R* = 1.2, our results suggest that the same testing regime with the largest 20% of households in an urban area is as effective as imposing strict lockdown measures and will curb the outbreak in a few weeks. Pooled household testing appears to be a powerful alternative to more invasive measures as a localized early response to contain epidemic outbreaks.

---

Protocols for testing the presence of COVID-19 were available (*1*) well before it was declared a pandemic by WHO on March 11, 2020 (*2*). In contrast, despite an unprecedented international effort to develop vaccines against SARS-Cov-2, the production launch of a successful vaccine has still not materialized. Due to time-consuming clinical trials that any vaccine candidate has to pass, this asymmetry is likely to characterize the majority of new virus pandemics that may emerge. Thus, exposure to a new virus pandemic is characterized by at least an 18 month window when accounting for the time lag between production launch and appearance of a substantial intervention effect from a vaccine (*3*). During this time, we are forced to curb spread through exploiting interventions provided by population testing and other non-pharmaceutical measures (*4*), including community, city and region-wide lock-downs.

The ability of a given test regime to inhibit spread has a substantial impact on the need for other measures: the greater the effect of testing, the fewer social restrictions need to be invoked, some of which carry huge societal costs compared to the expenses attached to testing. Thus, the identification of optimal test regimes in terms of efficacy, logistical feasibility and economic cost as a function of infection dynamics, deserves close attention. Here we show that, for any pandemic-causing virus behaving similar to SARS-CoV-2 (*5*), localized targeted testing of large households and subsequent quarantining of positive cases, is a highly efficient strategy for getting the infection dynamics under control.

Most countries have implemented a COVID-19 test regime targeting symptomatic cases combined with contact tracing. In addition to the substantial economic costs related to contact tracing, there are several inherent problems associated with this approach. First, the time from exposure to onset of symptoms is estimated to be 2-12 days, with a mean of 5.5 (*6*); Second, there is typically a multi-day lag after the first onset of symptoms until a test is performed; Third, voluntary symptomatic testing will leave many people with weak symptoms untested for a multitude of socioeconomic reasons (*7*); Finally, the obvious inability to systematically identify the considerable fraction of asymptomatic spreaders (*8–10*) implies that an important infection source is not targeted. Even with an exemplary implementation of symptomatic testing and contact tracing, a large number of the uncovered positive cases will have spent a significant portion of their infectious period unmitigated, and only a limited number of the potential disease-transmission chains in a society will be severed. This warrants the search for alternative feasible test regimes lacking the shortcomings of symptomatic testing.

In order to reduce the reproductive number *R* of a spreading process, network theory has shown that it is effective to immunize high-connectivity nodes (hubs) in heterogeneous systems (*11, 12*). Thus, the detection and isolation of hubs in social interaction networks can be considered the cousin of vaccination: while it does not prevent the primary infection, it does prevent secondary infections. Anticipating that private households are major hubs for the spread of COVID-19, we analyzed in-depth contact-tracing data available for 5, 531 of the laboratory confirmed cases occurring in Norway since the outbreak. The data confirm the most likely location for exposure to be private households (37.4%) (*13*). Furthermore, detailed demographic data from France’s National Institute of Statistics and Economic Studies (INSEE) (*14*) combined with epidemiological data from the French Agency for Public Health (Santé publique) (*15*) for the 96 European departments of France, show that household size is an important parameter in the spread of COVID-19 (Fig. 1). Calculating the correlation between available demographic variables and the extent of COVID-19 hospitalization, we find that the strongest correlates are dominated by household and household-related variables (Fig. 1A). In particular, the strongest correlated variable is the fraction of households with more than 4 persons (Fig. 1B). Similarly, the population shares of young adults (ages 20 − 39 years) and pre-school children are amongst the four strongest positive correlates, indicating a link between families with children and COVID-19 spreading. In contrast, the population share of older people (ages 60+ years) is strongly negatively correlated with the spread of the disease.

**Fig. 1.**
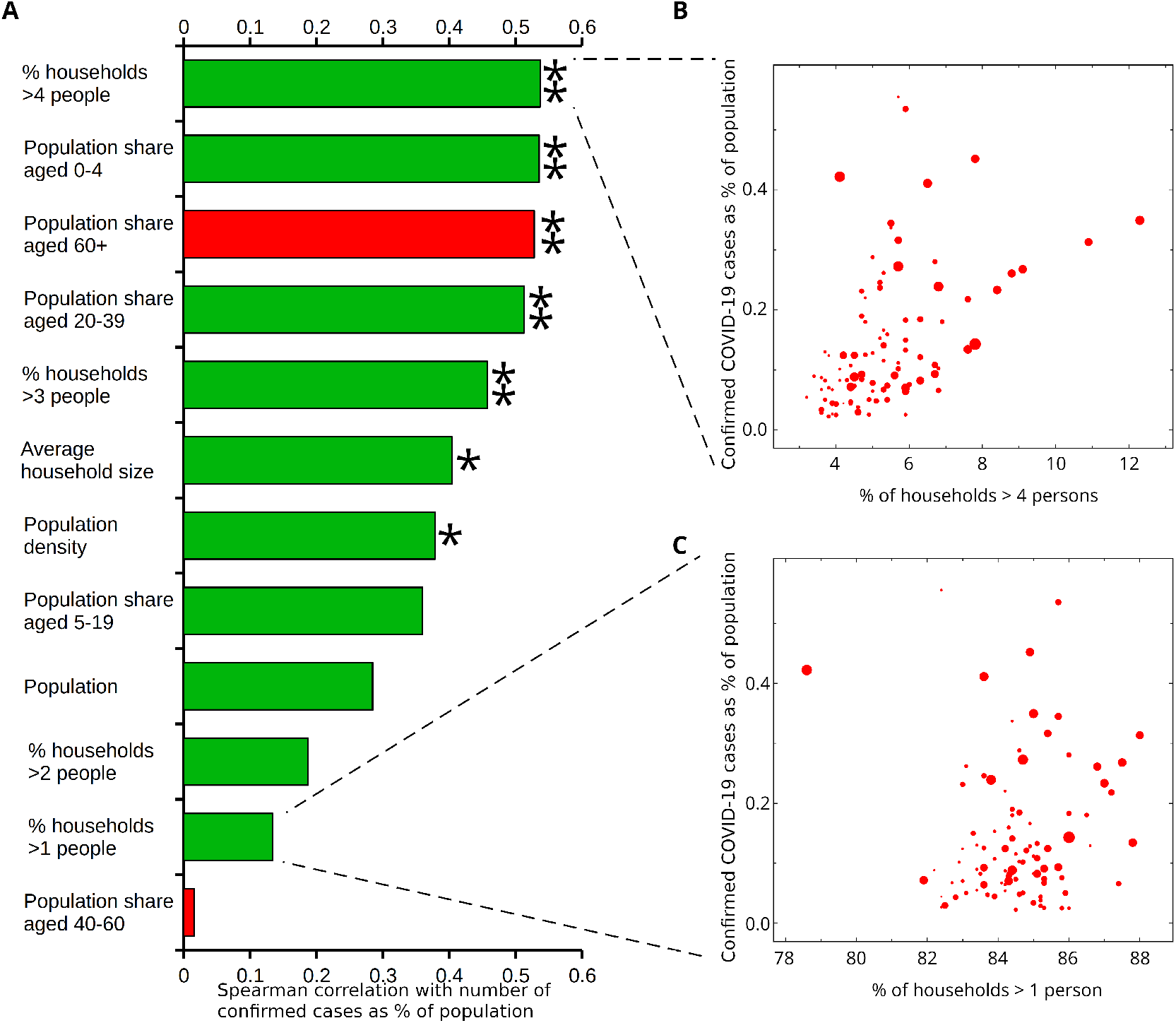
Demographic parameters associated with COVID-19 spread in France. (**A**) Correlation histogram (positive, green; negative, red) shows larger household sizes significantly correlated with levels of COVID-19 hospitalizations. Single (double) star indicates Bonferroni-corrected significance *P* < 0.01 (*P* < 0.001). Panels (**B**) and (**C**) show scatter plots of confirmed cases (as percentage of population) for the 96 departments of European France as function of the percentage of households larger than four persons and one, respectively. Size of markers is proportional to the population of each department.

The connection between large households and epidemic spread motivated us to test the intervention efficacy of a Targeted Pooled Household Testing (TPHT) strategy, defined by regular and scheduled pooled testing (*16*) of the largest households in a region followed by quarantining of the entire household upon a positive pooled test. To this end, we developed a high-fidelity individual-based model that uses detailed demographic data for each of Norway’s 356 municipalities (see Supplementary Text for details). The model consists of a multilayer network (*17*), in which each layer represents a type of interpersonal relationship (such as household, work, education, and random encounters) with an associated infectivity. Using hospitalization data for Oslo (*18*) during the 2.5-month period from March 1st to May 15th as a target (Fig. 2A, red line), we calibrated the infectivity of each layer by subjecting the model to the same course of shutdowns and re-openings as those mandated by Norwegian authorities in the same period. Not only is the model able to obtain a close fit with the hospitalization data used for calibration, but the parameters identified also prove predictive within confidence intervals for the period from mid-May to at least late August (Fig. 2A, green line). In particular, the identified parameters (Supplementary Table 1) correspond to an expected reproductive number of *R* ≈ 1.05 for the period from May 15th until the end of June.

**Fig. 2.**
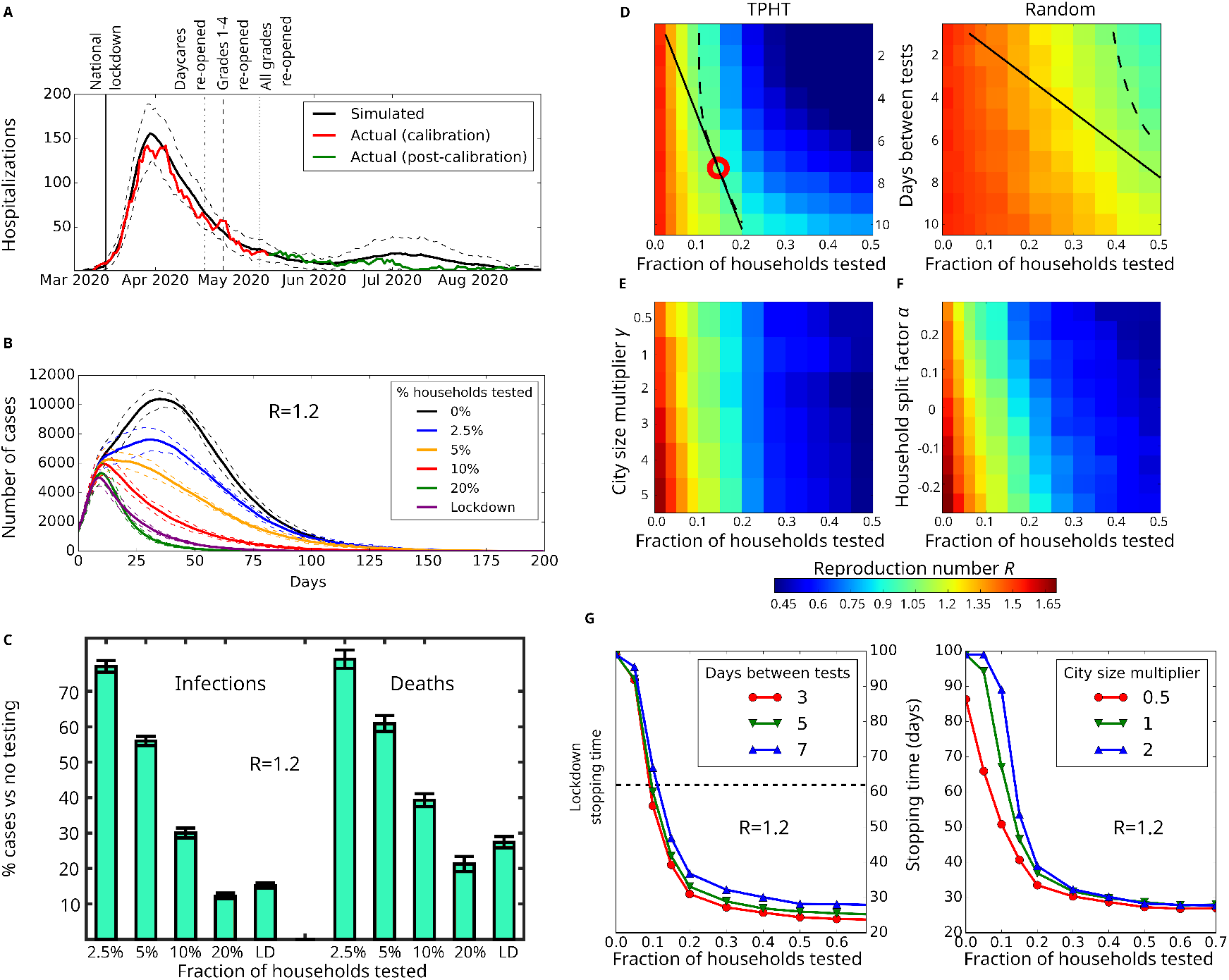
Simulation results and effectiveness of targeted pooled household testing (TPHT) on COVID-19. (**A**) Fitting the model to Oslo hospitalization data. We plot the mean predicted number of hospitalizations (black) and confidence interval (2*σ*, dashed). Actual Oslo hospitalizations (red) were used as calibration (until May 15th). Hospitalization data May 15th-August 30th (green) were not used to determine model parameters. (**B**) Effect of TPHT in response to a sudden rise in cases (reaching 1,000 symptomatic individuals), assuming general infectivity parameters similar to those of Oslo in late May 2020 but with 50% increased infectivity of random contacts giving *R* = 1.2. (**C**) Predicted number of deaths and infections for different TPHT testing fractions corresponding to panel (**B**), relative to no testing. Panels (**D**)-(**F**) use same parameters as panel (**B**), except with a 165% increased infectivity of random contacts giving *R* = 1.5. (**D**) Effect of test frequency and fraction on *R*, for TPHT (left) and random pooled household testing (right). Dashed and solid lines indicate isoclines for *R* = 1 and constant test density, respectively. Optimal point is marked with red circle. (**E**) Response of weekly TPHT to varying city size. We scale the population of the baseline Oslo model (*γ* = 1) to generate larger (*γ >* 1) or smaller networks with household, school, daycare, workplace and nursing home size-distributions unchanged. (**F**) Response of weekly TPHT to changes in distribution of household size, relative to the baseline Oslo model (*α* = 0). For *α >* 0, a portion of households are each split into a new pair, yielding a smaller average-household size than the baseline. For *α* < 0, a portion of the households are pairwise merged, yielding a larger average household size than the baseline. Household, school, daycare, workplace and nursing home size-distributions are kept unchanged. (**G**) COVID-19 stopping time (number of days until symptomatic cases are reduced by 75%) in response to changing days between tests (left) and fraction of weekly TPHT tests (right). Stopping times longer than 100 days are truncated.

Assured by the above validation results, we investigated the ability of weekly TPHT to contain an outbreak of 1, 000 symptomatic cases in Oslo, a city with ∼ 700, 000 inhabitants (see Supplementary Text for details). We tested the infection spread situation with *R* = 1.2 (Fig. 2B), obtained by increasing the chance of infection in the random contact layer by 50%. This is a little more than the estimated *R* = 1.1 value for Oslo in September, 2020 (*19*). We find that a TPHT involving a mere 2.5% of the households is able to noticeably reduce the number of cases (see Supplementary text for details on TPHT selection). A weekly 10% TPHT-level reduces the number of infected by ∼ 70% and dead by 60% relative to no testing (Fig. 2C). Using less than 20% weekly TPHT, the intervention effect is on par or better (Fig. 2C) than what was achieved by the national lockdown regime (Fig. 2B, purple) imposed in Norway from March 13th until April 20th (Fig. 2A). It should be noted that a 10% TPHT-level corresponds to testing only about 5% of the Oslo population weekly.

Assuming a situation with an *R* = 1.5, similar to the experience of many European countries during fall 2020 (*20*), we then explored the ability of TPHT to reduce *R* by varying the frequency and fraction of households to be tested (Fig. 2D). We determined the optimal balance between frequency of testing and number of tests needed to achieve the critical point *R* = 1 by finding the density of daily TPHT that intersects the *R* = 1 isocline to be *ρ* = *x/y* = 0.05, with *x* being the fraction of TPHT performed per *y*-day interval. The model predicts that the optimal allocation to contain the epidemic with *R* = 1.5 is to implement a testing regime where the 15% largest households are tested every seven days (see Supplementary text, “Description of the TPHT process”, for details), corresponding to a number of tests equalling 2.14% of the population every day. The necessary density of daily tests is significantly reduced for smaller *R* (see Supplementary Figure 1 for *R* = 1.2). In comparison, we find that regular pooled testing of randomly selected households would require an infeasible number of tests to obtain an appreciable reduction in *R* (Fig. 2D). This illustrates that the prioritization of larger households in TPHT leads to a dramatic impact on *R*, allowing for an effective suppression of infection spikes with a moderately ambitious weekly test capacity (Fig. 2B).

When faced with an outbreak, its estimated duration is an important parameter for health authorities when planning mitigation. We define the outbreak stopping time as the duration until the outbreak is at a 25% level relative to when interventions were implemented. First we computed the stopping time associated with the Norwegian national lockdown regime in effect from March 13th (Fig. 2A), by extending the lockdown indefinitely without further re-openings. We find that this situation corresponds to a stopping time of 61 ± 3 days, in agreement with the 60 days observed in public prevalence data (*18*). For the outbreak in Fig. 2B, we measured the stopping time as function of TPHT fraction for different test frequencies (Fig. 2G, left panel). The majority of cases giving the lowest stopping times is achieved by TPHT fractions in the range 0.20 − 0.25, regardless of test frequency. For instance, weekly testing of the top 20% largest households (≈ 2.86% per day) achieves a stopping time of 36 days, which is faster than test regimes of 15% of households every 5 days (stopping time of 42 days) or 10% of households every 3 days (stopping time of 55 days), both of which demand a higher combined testing capacity (3% per day and ≈ 3.3% per day).

To assess the generality of our results to other population regions, we investigated the effect of independently varying the population size (Fig. 2F) and the household size-distribution (Fig. 2G) while conducting weekly TPHT. Using a population multiplier (*γ*), we scaled the size of our base-line model (*γ* = 1) while keeping all other demographic distributions unchanged. We find that larger cities need a larger TPHT fraction in order to bring *R* down to the same amount as in a smaller city. A likely explanation for this is the presence of scale-free spreading dynamics in our model, the impact of which has previously been shown to increase with larger network sizes (*21*). The stopping time for weekly TPHT in three different population sizes (Fig. 2G, right) drops rapidly with increased TPHT test fraction until reaching a value of 0.3, for which it has achieved near saturation. To investigate how changes to the household-size distribution impact the ability of TPHT to curb spread, we introduced a household-split factor *α* that determines if a pair of households should be joined (*α* < 0) or a single household should be split into two (*α >* 0), while keeping the population size unchanged (see Supplementary Text for details). As anticipated, we find that *R* increases with the proportion of large households (Fig. 2F). By increasing the weekly test fraction, one may countermand this effect, and application of TPHT is still capable of reducing *R* to below unity (Fig. 2F). Thus, even though the demographic characteristics of a city has a noticeable impact on *R*, figures 2D-2G indicate that TPHT is able to drive *R* below unity for modest test fractions across a variety of city sizes and household compositions.

In summary, we have found that the implementation of a TPHT regime is broadly effective in breaking infection chains before their spread potential has been realized. As a TPHT regime can be combined with a wide range of economically benign social distancing measures, our results suggest that it represents an attractive option for bringing a pandemic outbreak under control without inducing the societal costs of large-scale quarantines, lock-downs and curfews.

## Data Availability

The code used for simulations included in the manuscript is available at DOI:
10.6084/m9.figshare.13143575
Further developments of the main model are available at https://github.com/andrevo/covid19-ntnu

## Acknowledgements

We thank Profs. Thor Inge Fossen, Morten Hovd, and Ingelin Steinsland for comments and feedback, and HUNT Cloud (https://www.ntnu.edu/mh/huntcloud) for computing resources.

## Author contributions

AV, SWO and EA conceived the project. AV built the software with support from NM, EK, MH, and KN. AV, NM and EA analyzed the data. AV, SWO, NM and EA wrote the paper.

## Data and materials availability

All data are available in the manuscript or the supplementary materials.

