## Supplementary material for "Containing pandemics through targeted testing of households"

### Supplementary Text

#### Description of individual-based modeling (IBM) software

When selecting and designing our computational modeling framework, we have explicitly taken into consideration the following aspects: (A) It should be possible to directly test a wide variety of interventions, such as partial workplace and school closings, social distancing, and testing with subsequent isolation of infected individuals. (B) Each infected person should have a disease-state development where the stages have realistic time delays. If a lag in response time is not included, we anticipate that all estimated response times to interventions will be markedly wrong. This is especially evident for the correct estimation of R, and thus, the capacity to determine how quickly R responds to interventions. The mentioned challenges are inherent to most metapopulation-based models, whereas an IBM approach is not hampered with such issues.

Guided by the above reasoning, we have developed a complex system modelling framework to describe the spread pattern of COVID-19 in mainland Norway. The model design and build features are chosen with the intent that it can be used to carefully assess a multitude of relevant intervention strategies. Briefly, the model is based on using an IBM based on complex network theory for each Norwegian municipality. The features of the model’s layers and each individual all adhere to high-resolution demographic data specific for the respective municipality. The disease-state of each individual follows an epidemiological SEIR-type model.

Our total model for all combined municipalities contains approximately 5.3 million people and simulates dynamics of spread through social interactions in layers such as households, schools and workplaces.

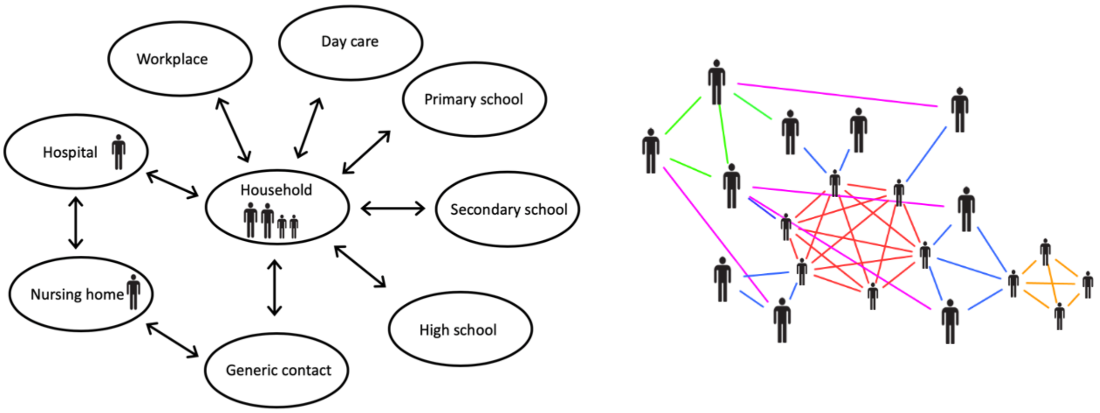

*Fig. 1* Structure of network IBM. (Left) Possible domicile indicated by person figure. Named circles show available layers that a person can be member of. (Right) Illustrative example of resulting connection network between individuals caused by shared group membership in different layers: household (blue), primary school (red), day care (orange), work (green) and generic (pink).

### Stochastic IBM network with SEIR epidemiological transmission dynamics

#### Structure of the municipality IBM network

We generate a high-fidelity IBM for a single municipality by creating a set of households corresponding to the population $N_m$ of that municipality. Each household consists of one or several nodes (people), to each of which we assign a list of attributes:

1. Age
2. Domicile
3. Layer memberships / group membership within layers (see Fig. 1)
4. Disease state (see Fig. 2), and date of last change in disease state
5. Disease test state

The model has 9 layers. Layer a)-h) each consist of many groups, and each individual is only member of one of these groups. Further, an individual can only be present in a single of the layers b)-h). The layers are: a) Household, b) Day-care, c) Primary school, d) Secondary school, e) High school, f) Workplace, g) Nursing home, h) Hospital, and i) Generic contact network. A group is designed as a k-clique, i.e. where all

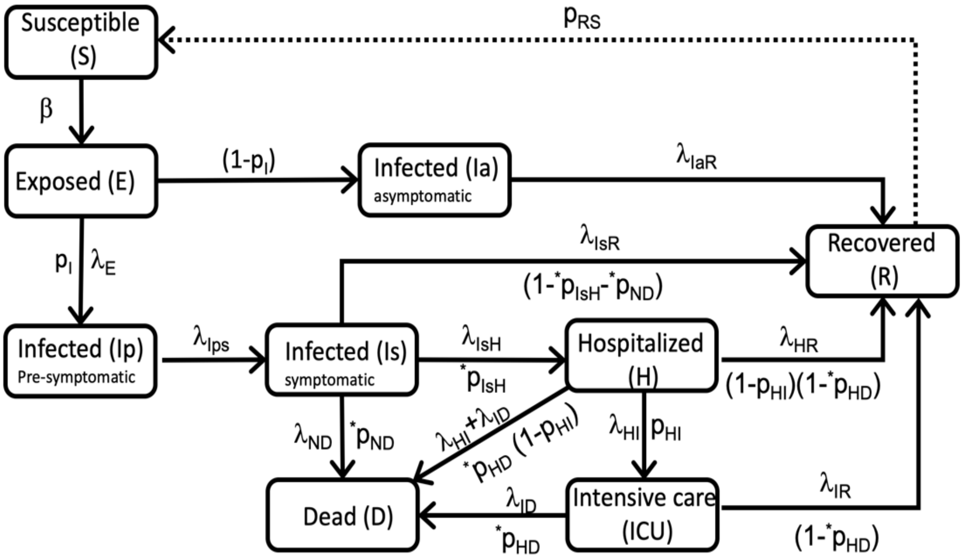

*Fig. 2* Block diagram showing schematic of the SEIR-type epidemiological state model for each individual in the IBM. Solid lines show transitions currently active in the model. The dotted line indicates current uncertainty about long-term COVID-19 immunity. Starred parameters indicate age dependent values.

members of a group are in contact. The assignments that form the IBM contact network are constrained such that known high-resolution demographic data for each municipality is matched:

1) The household layer consists of separate households with a size and age distributions that follow demographic data for that municipality.
2) The number of schools, type, and their student populations are based on demographic data.
3) The number of day care facilities is based on demographic data.
4) The number of nursing homes and their population sizes are based on demographic data.
5) If a household has multiple children in e.g. day-care age, these children are assigned the same day-care. Similar for primary and secondary schools. For high-school age, the assignment to school unit is random.
6) In the work layer, the number of companies and their sizes are based on demographic data. Note that this layer is intended to represent spread between co-workers. For professions with large amounts of exposure to the general public, contacts with customers are represented through the generic contact network.
7) The generic contact layer is designed to capture heterogeneity in a person’s daily contact patterns. While the other layers consist of fixed cliques, in the random layer, each node

connects to a random selection of other nodes for each simulated day. For each connection between an infected and a susceptible node, there is a fixed probability of the susceptible node becoming infected. The daily number of connections for each node is redrawn each day, according to a uniform distribution ranging from 1 to a maximum number of daily contacts $C_D$, which is a node attribute assigned at the initiation of the simulation.

8) We include two different modes: (A) For young (< 20) and elderly (80+) we assign a maximum number of daily contacts $C_D$ following a normal distribution, whereas the remaining age groups are assigned contacts following a combination of a normal and a power law distribution: Contact number is drawn from (i) normal distribution, (ii) power law before being added together. The latter generates larger contact heterogeneity.
9) Using data from Statistics Norway[1], adults with different work and household municipalities are assigned work locations in the correct municipality. Supplementary Table 2 lists the data tables used.
10) When a person is committed to a hospital, they are removed from their domicile (household or nursing home).
11) When an infected person manifests a symptom, is confirmed COVID-19 positive, or an asymptomatic person is confirmed COVID-19 positive, the model assumes they will self-quarantine from activities in all layers except their domicile.

All individuals participate in the generic contact network, which is designed as a random time-dependent scale-free network to capture heterogeneity in contact patterns. A new instance of this network (per municipality) is generated new every day. Except for the generic contact network, each grouping in each layer is represented as a k-clique, where every individual is connected to all others in the group, thus representing a well-mixed group. Fig. 1 shows a schematic of the IBM layers inside

[1] Statistisk Sentralbyrå (SSB). https://www.ssb.no/statbank/

which there are smaller groups (left panel) and a simplified example of a possible resulting contact network (right panel). The probability of infection depends on which layer the infected contact is located. Within a layer, all members of a single group have a constant probability of infecting other members of that group. The specific values used for the parameters in each layer are detailed in Supplementary Table 1.

#### Population size scaling

We developed a schema for scaling up the population size of a municipality that preserves its demographic distributions. The scale parameter $\gamma$ determines the new size of a municipality: $\gamma$ =1 for the unscaled population, $\gamma$ = 0.5 results in a municipality with half of the population, and $\gamma$ =2 gives a municipality with twice the original population. As the statistical data, age data and household-, nursing home- and workplace-compositions obtained from Statistics Norway take the form of percentages, the distributions are independent of the size of the network and can be used as provided, simply by multiplying by $\gamma$ the number of desired individuals/households/workplaces (provided by Statistics Norway and passed directly as inputs to the generating code when $\gamma = 1$). On the other hand, schools and daycares are explicitly and individually defined, and their numbers are too few to be mapped to fine-grained size distribution. Therefore, in order to scale these systems to match the scaling of the whole municipality, we instead opt for an approach where we randomly select and duplicate (or erase, for $\gamma < 1$) schools and daycares in the original municipality data until we reach the number specified by the scale factor.

#### Variation of household-size distribution

In order to evaluate the impact of the household size distribution on spreading potential, we needed to establish a systematic way to modify the distribution, preferably defined by a single scalable parameter α. We decided on either splitting or merging existing a fraction of households in the original distribution, yielding smaller or larger households respectively. In the first case, we identify households eligible for splitting, using the criteria that it must contain at least two adults. This is necessary in order to ensure that we do not create households containing only children. We then define α as the fraction of eligible households to split. The split itself is conducted by assigning the two first adults in the selected household to their own household, with each of the remaining individuals assigned to either household by coin flip (Bernoulli, 0.5).

We represent the merging of households by using negative values of α: As merging households with at least one adult each will naturally lead to a new household with at least one adult, all households are eligible for merging. We therefore randomly assign the households into pairs, with a chance |α| that determines the fractions of pairs which are combined to form a single household consisting of the members of the two original households, reducing the total number of households in the system by one for each merged pair. The households in the remaining pairs stay unchanged.
As an example, consider a fictitious small municipality containing 2000 individuals, labeled *i1, i2*, *i3*, … , distributed across 1000 households *H1, H2, H3, …* such that *H1* contains individuals *i1, i2* (using the notation *H1 = [i1, i2], H2 = [i3]* (a lone individual), *H3 = [i4, i5, i6, i7]* , and *H4 = [i8, i9].* We then pair household H1 with household H2 and household H3 with household H4 and label this list *X*, so that *X1 = [H1, H2] = [i1, i2, i3], X2 = [H3, H4] = [i4, i5, i6, i7, i8, i9], ….* Our next step is to initiate an empty list of new households, which we label *N*. We take *X1,* and with probability |α| add *N1 = [i1, i2, i3],* otherwise we add *N1 = H1 = [i1, i2]* and *N2 = H2 = [i3]*. We repeat this process for *X2,* adding *H3* and *H4* to *N* either separately (with probability 1-|α|) or as one combined household *[i4, i5, i6, i7, i8, i9]* (probability |α|), and similarly for each pair in X. If the number of households is odd, one household is copied directly into *N* without possibility of pairing. The remaining households are assigned to pairs as described.

Note that *N* will have a lower total number of households than *H*, but the same total number of individuals. For instance, setting α = 0.1 in a network starting with 1000 households, the average result is *N* consisting of 950 households (900 households in 450 pairs remain unchanged, with the remaining 100 households merged pairwise into 50 new ones). For α = 0.5 in a network starting with 1000 households, we would get (on average) 750 households, with 500 households unchanged and the other 500 being combined pairwise in order to form 250 households.

#### SEIR-type epidemiological dynamics

Each individual in the IBM model is either healthy, in various states of infection, or recovered from the disease: Susceptible (S); Exposed (E); Infected, asymptomatic (Ia); Infected pre-symptomatic (Ip); Infected, symptomatic (Is); Hospitalized (H); Intensive care (ICU); Recovered (R); or Dead (D). The different states and their possible transitions are captured by the state-transition schematic of Fig. 2. The direct transition from Is to D captures the disease trajectory for some people in nursing homes. The transition from S to E is governed by disease transmission probabilities in the IBM contact network. After an individual is infected from a neighbor in the contact network, the SEIR-type dynamics of that individual’s disease state (onward from E) are governed by stochastic processes with appropriate waiting times according to empirical data for COVID-19. We continually update our parameter estimations given the daily update in disease state numbers for Norway by region.

At each time point, an individual will store four points of information about their progression through the SEIR states: current state, date of last change of state, next state, and date of next change of state. Each day, the model updates the state as needed. Upon entering a new state, the model selects the following state according to the probabilities and waiting times specified in Fig. 2 and Table 1. Once the next state is determined, the duration for which the individual will remain in the recently entered state is generated according to that state’s specific λ, and the date of next change is set accordingly. Note that our SEIR-model uses age-stratified transition rates, where the age of an individual decides which of the age groups that individual is part of. Supplementary Table 1 shows the rates used for the different SEIR transitions of Fig. 2.

#### Implementation of intervention measures

A high-fidelity IBM allows for the implementation of detailed interventions, where contact patterns of a selection, or all, of the individuals may be modified. It is thus straightforward to conduct computational experiments where some or all schools are closed or partially closed, various levels of social distancing are included, or testing and vaccination is administered. The most basic intervention strategy consists in locking down various IBM-layers. How this is implemented in detail depends on the layer:

1. For day care facilities, the layer is disabled in its entirety. Similarly for secondary schools and high schools.
2. For primary schools, a shutdown is implemented separately for grades 1.-4. and grades 5.-7., allowing for the opening of grades 1.-4. independent of the other grades. The motivation for this is to be able to mimic actual interventions imposed by the Norwegian authorities.
3. In the work layer, we implement a gradual shutdown by specifying that a fraction of cliques, intended to represent workplaces where work-from-home is feasible, are disabled.
4. In the random contact layer, lockdown is implemented by setting an upper bound on the number of possible daily contacts for a fraction of the nodes.
5. The household and nursing home layers are never disabled.

General social distancing and hygiene measures are represented by a scale factor multiplied with the base transmission rates of each layer.

#### Targeted quarantine of individuals

The quarantining of an individual is represented by disabling workplace, school, and random layer spread for asymptomatic and pre-symptomatic individuals put into quarantine, similarly to behavior patterns for individuals who self-quarantine when symptoms manifest. Such quarantines are imposed on individuals who test positive, as well as other individuals at high risk of contagion from someone who has recently tested positive (such as those in the same household, workplace, or school).

##### COVID-19 testing of (sub-) population

The testing itself is represented by drawing individuals from the symptom-free population (susceptible, latent, asymptomatic, pre-symptomatic, or recovered), and returning a positive test if they are asymptomatic or pre-symptomatic. Pooled tests are represented similarly, but with multiple people (forming a pool) simultaneously tested, and all tests marked as positive if any of the people in the test pool are in the asymptomatic or pre-symptomatic states. Our IBM implementation allows for random selection of individuals for testing, or testing based on a pre-conceived schema for node attributes.

#### Base-line fitting of IBM to empirical data

The parameters in the stochastic IBM are treated as fixed, and they are either set from literature or estimated directly from publicly available empirical data, as indicated in Supplementary Table 1. We determined model parameters by fitting the predicted hospitalization rate of our Oslo model to hospitalizations for Oslo in the period from March 1st to April 20th. We assume a sudden transition between two regimes, with infections following original “unrestricted” probabilities until March 13th. The “unrestricted” regime is initialized with 20 cases at an unspecified date and run until the system reaches $N_{lock}$= 400 active cases. This point is marked as March 13th, and serves as a reference point for the remainder of the simulation. The motivation for fixing dates at this point (rather than the initial point of 20 cases) is two-fold: First, it is important that the model follows a progression of non-pharmaceutical interventions (NPIs) that actually corresponds to those implemented by Norwegian authorities. Therefore, we decided to use the earliest NPIs introduced (March 13th) as an anchoring point, with the dates of subsequent NPIs defined in relation to the first. Second, due to the heterogeneity in the infectious potential of individuals (with a small fraction of individuals being substantially more infectious than others due to their particular contact network), the model exhibits highly stochastic behavior when the number of infections is low, as the number of high-infectivity individuals infected is more variable. As small fluctuations in the early evolution of the epidemic can have dramatic effects on long-term forecasts, anchoring the network around a reasonably large number of cases allows the model to make more consistent predictions. This approach also solves the issue of highly stochastic behavior at low infection levels, as 400 active cases is sufficient to reduce this notably.

After these steps, all schools and day care facilities close, as do 50% of workplaces. Random contacts are reduced by 83%. Infection probabilities in nursing homes and within-households remain the same in both regimes. Model simulations start in March and continue until August 30$^{th}$, 2020.

Since the lockdown in Norway occurred in the early stages of the epidemic, the fitting of the IBM results to data depends very much on the number of symptomatic infected on the day of lock down (March 13, 2020). Thus, for a given parameter set and municipality, we have used a large number of simulations to determine the optimal number of infected ($N_{lock}$) on this day for the model predictions to fit to time series data of number of hospitalized persons. Simulations are subsequently conducted by initiating the network with a small number of infections and running the code until $N_{lock}$ is reached. As described above, the date is set to be March 13 at this point. We anticipate that several of the IBM parameters can be estimated from other data sources when they become available, e.g. large-scale systematic testing regimes for COVID-19.

#### Description of the TPHT process

##### Selection of households

The central aspect of TPHT is the prioritizing households for testing according to size. Consider a municipality with the following distribution of households:

- 6 persons and over: 3 thousand households
- 5 persons: 7 thousand households
- 4 persons: 30 thousand households
- 3 persons: 20 thousand households
- 2 persons: 50 thousand households
- 1 person: 90 thousand households.
- Total: 200 thousand households, population of above 420 thousand (depending on the exact distribution of households of size 5 and more)

In this case, a TPHT test fraction of 2.5% corresponds to 5 thousand households, meaning we select all households 6 and over (3 thousand), and a random choice of 2 thousand of the 5-person households. A TPHT test fraction of 5% would yield 10 thousand households, meaning all households 5 and above. A TPHT test fraction of 10% gives 20 000 households, meaning all 10 thousand households five and above, as well as 10 thousand randomly chosen 4-person households. For a given fraction and test intervals, the selected households are distributed evenly across the interval. For instance, using a test fraction of 3.5% on the municipality above, with a test interval of 7 days, we would have to test 7 thousand households a week; with one thousand periodically tested every Monday, another thousand tested every Tuesday, and so on, for the duration of the TPHT test regime.

#### Pooled household testing and quarantine

Pooled testing is a method by which samples from multiple individuals (a pool) are combined and jointly analyzed, returning a positive if any of the individuals constituting the pool are infected, without giving further information as to *which* of the individuals in the pool are infected (*Sunjaya & Sunjaya*, 2020).

Due to the household constituting a high-risk environment for transmission that is unaffected by many of the interventions introduced to curb epidemic spread (such as work from home, school shutdowns, closing of public venues and so on), TPHT proposes treating each household as a common unit with regards to testing of isolation. In short, if one household member is confirmed to be sick, all household members should be treated as if they were, preventing the infection from being passed on through society at large through undiagnosed family members.

#### Estimation of R for a choice of simulation parameters

In order to estimate the reproductive number *R* associated with a given parameter set, we begin by noting that our individuals have a time-independent infection profile, i.e., a given individual is equally infectious every day of the disease (past the initial latent phase). It follows that the daily number of new infections per already sick individual equals R divided by the typical disease duration of 9 days. Reversing this, we can estimate R at a given time point by taking the daily number of new infections, divided by the number of pre-existing cases for that day (yielding the daily infection rate per person), and multiplying that by 9 days.

To illustrate this, we use this approach to calculate R for a single simulation run as in Fig 2B. Setting $t=1$ day, we can compute an effective daily R, as shown below for an initial period of 10 days with infectivity parameters set to pre-lockdown values (early March), after which we abruptly transition to a regime corresponding to late May.

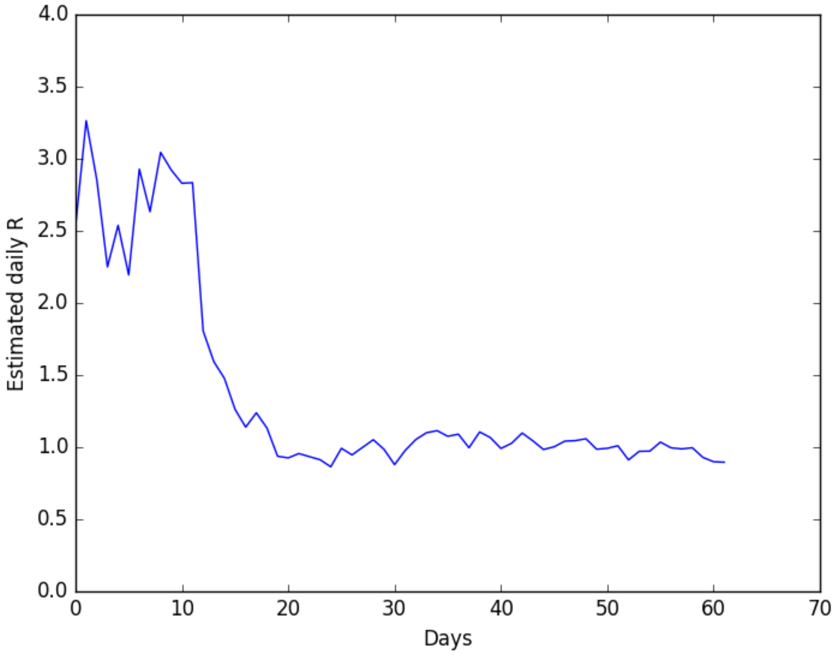

*Fig. 3:* Daily estimated R for a test regime consisting of 10 days with early March parameters, with an abrupt transition to late May parameters at *t*=10 days and on.

We make two key observations: first, there is some stochasticity in the determined value of *R* between consecutive days; second, the transition from one regime to the next exhibits an intermediary phase of a few days until *R* begins to stabilize around a new average. Meanwhile, estimating R over longer periods of time leads to substantial discrepancies: if R > 1, there is a build-up of immunity, leading to a gradual reduction in R; if R < 1, infection levels will gradually drop off, leading to situations with very few infected and correspondingly high noise in the relative number of new daily infections. Because of this, for purposes of determining R, our process is as follows:

- First, initialize the model with 20 infected individuals
- Unfettered growth until 100 symptomatic cases are reached (this is enough to remove most of the noise, while not enough to build up substantial immunity in the population). The purpose of this approach (rather than just beginning with a 100 infected individuals on the first day) is described in the next section.

- Once 100 (simultaneous) symptomatic cases are reached, transition into the regime for which we wish to determine R. Due to the high infectivity up to this point, there is also a substantial number of pre-symptomatic cases already infected at this point.
- Take the daily average R from 12 to 17 days after the transition mentioned above (equivalent to the 22-27 day interval in Fig. 3 above). For Fig. 2D-F in the main article, we perform 20 duplicate simulations per color point, for FIg. 2B, C and G, we perform 40 replicate simulations per data point.

#### Simulation of outbreaks

When conducting the simulation of an outbreak consisting of $N_{out}$=1,000 infected. the simple approach is to randomly select $N_{out}$ nodes in the network and abruptly change their state from susceptible to exposed. However, this will cause artificial oscillations in the data since all of the exposed individuals start out in synchrony because they were infected at the same time. It takes several infection generations for these oscillations to subside. Instead, we dynamically generate $N_{out}$ by starting from a small seed of 20 randomly (simultaneously) selected and exposed individuals. Subsequently, the IBM is simulated until we reach $N_{out}$ symptomatic individuals. This state is used as the starting point (day 0) of the outbreak simulation, from which interventions may be implemented. We go through this initiation process independently for each separate simulation, thus ensuring stochastic variation in the day 0 state.

#### Software availability

The IBM software used for performing the simulations is available on two general access platforms:

1. Active development on GitHub: https://github.com/andrevo/covid19-ntnu
2. Code snapshot used to generate the results of this publication are available on figshare at DOI: 10.6084/m9.figshare.13143575

### Supplementary Figure 1. TPHT for effect on R as function of test frequency and fraction with basal R=1.2

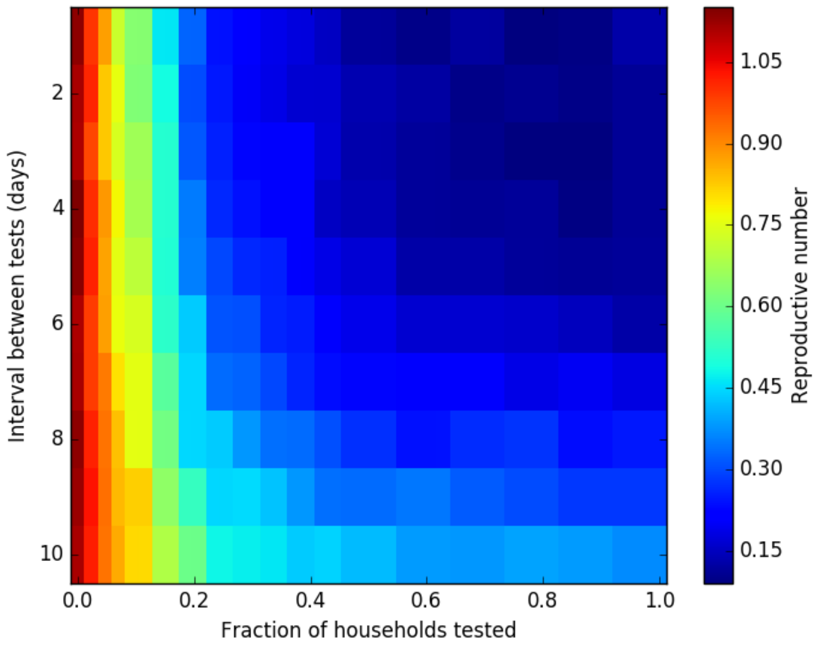

**Caption:** Effect of varying test frequency and test fraction on the reproduction number, assuming *R*=1.2 as the value without testing.

### Supplementary Figure 2. Household-size histograms.

When varying the household-size composition in the model, we used the approach detailed above in this document. Here, we show three of the resulting household-size histograms for $\alpha$=-0.3, 0, 0.3. Note that the bars sum up to 700,000 for each of the $\alpha$–values.

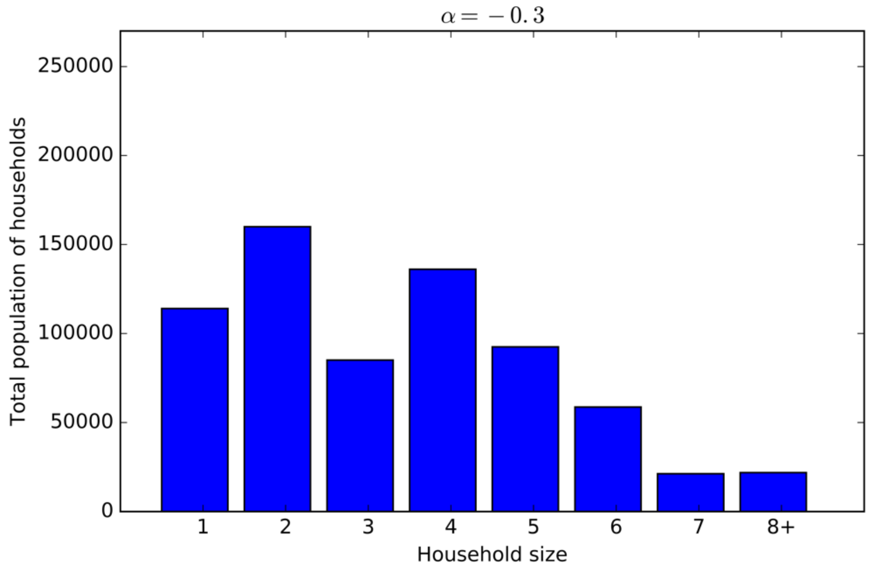

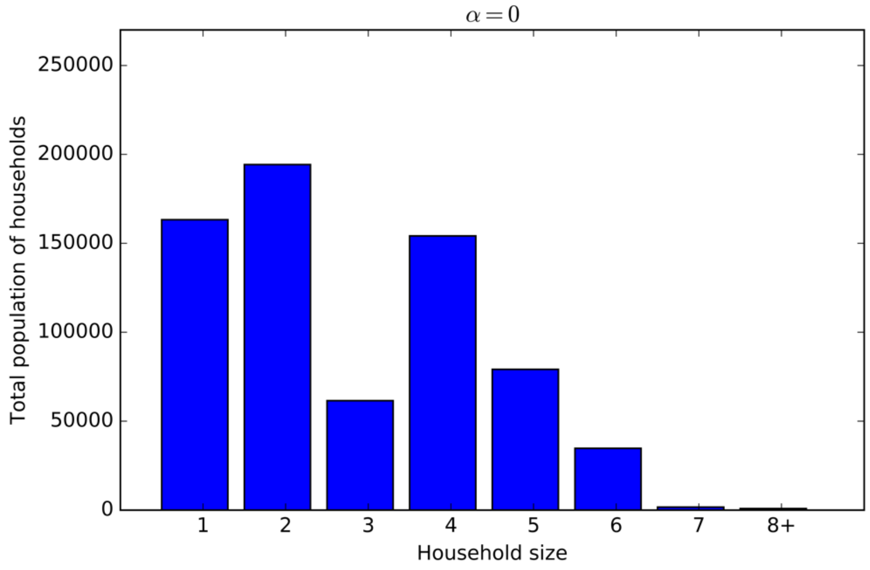

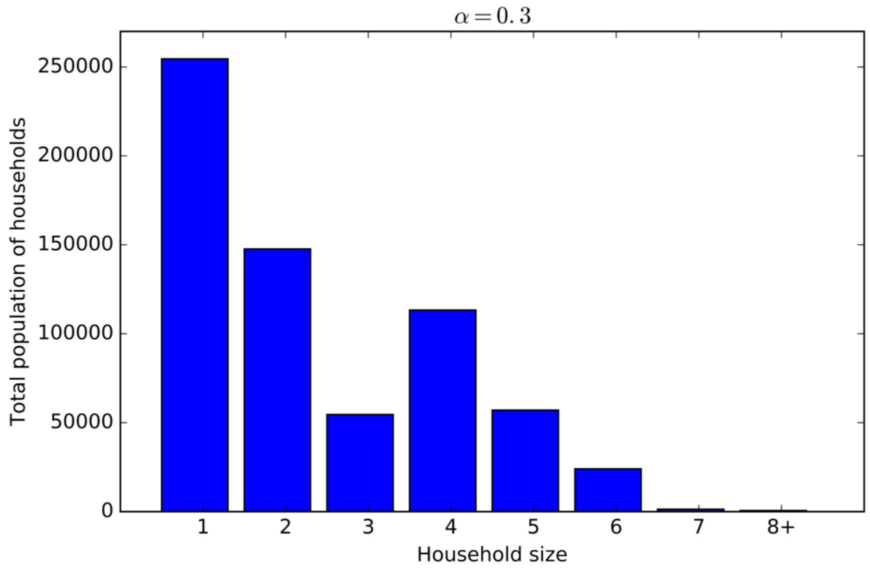

$\alpha = 0.3$
Total population of households
250000
200000
150000
100000
50000
0
1
2
3
4
5
6
7
8+
Household size

### Supplementary Table 1. Parameters used in simulations

| Model parameters | Symbol | Value | Function | Source |
|---|---|---|---|---|
| **SEIR-epidemics** | | | | |
| Probability of infection | b | ---- | Network effect |  |
| Days incubation time | $l_E$ | 1 | Fixed | |
| Days spent pre-symptomatic | $l_{Ips}$ | 5 | Poisson | Norwegian Institute of Public Health (FHI) |
| Days symptomatic before recovery | $l_{IsR}$ | 5 | Poisson | Norwegian Institute of Public Health (FHI) |
| Days symptomatic before hospitalization | $l_{IsH}$ | 6 | Poisson | Data from Health Region South East, Norway |
| Days symptomatic in nursing home before death | $l_{ND}$ | 10 | Poisson | Data from Health Region South East, Norway |
| Days in hospital before recovery (no ICU) | $l_{HR}$ | 8 | Poisson | Data from Health Region South East, Norway |
| Days in hospital before ICU | $l_{HI}$ | 4 | Poisson | Data from Health Region South East, Norway |
| Days in ICU before recovery | $l_{IR}$ | 12 | Poisson | Data from Health Region South East, Norway |
| Days in ICU before death | $l_{ID}$ | 12 | Poisson | Data from Health Region South East, Norway |
| Days asymptomatic before recovery | $l_{IaR}$ | 8 | Poisson | Norwegian Institute of Public Health (FHI) |
| % exposed developing symptoms | $P_I$ | 50 | Bernoulli | |
| % symptomatic dying outside of hospital: | $P_{ND}$ | | Bernoulli | |
| nursing home residents 70-79 years | | 26 | | Adjusted to Norwegian hosp. death rates, Verity et al |
| nursing home residents 80-89 years | | 42 | | Adjusted to Norwegian hosp. death rates, Verity et al |
| All others | | 0 | | |
| % hospitalized dying: | $P_{HD}$ | | Bernoulli | Verity et al, Lancet, 2020 |
| 0-9 years | | 1.61 e-3 | | |
| 10-19 years | | 6.95 e-3 | | |
| 20-29 years | | 3.09 e-2 | | |
| 30-39 years | | 8.44 e-2 | | |
| 40-49 years | | 0.161 | | |
| 50-59 years | | 0.595 | | |
| 60-69 years | | 1.93 | | |
| 70-79 years | | 4.28 | | |
| 80+ years | | 7.8 | | |
| % symptomatic being hospitalized | $P_{IsH}$ | | Bernoulli | Verity et al, Lancet, 2020 |
| 0-9 years | | 0 | | |
| 10-19 years | | 0.048 | | |
| 20-29 years | | 1.04 | | |
| 30-39 years | | 3.43 | | |

| | | | | |
|---|---|---|---|---|
| 40-49 years | | 4.25 | | |
| 50-59 years | | 8.16 | | |
| 60-69 years | | 11.8 | | |
| 70-79 years | | 16.6 | | |
| 80+ years | | 18.4 | | |
| % hospitalized needing ICU | $P_{HI}$ | 30 | Bernoulli | Fitted to Norwegian Institute of Public Health, ICU numbers |
| % not developing immunity | $P_{RS}$ | 0 | Bernoulli | |
| **Individual-based network model** | | | | |
| Infectiousness in Day Care | | 0.015% | Bernoulli | estimated IBM fit to Norwegian clinical data |
| Infectiousness in Primary School | | 0.002% | Bernoulli | estimated IBM fit to Norwegian clinical data |
| Infectiousness Secondary School | | 0.015% | Bernoulli | estimated IBM fit to Norwegian clinical data |
| Infectousness High Scool | | 0.015% | Bernoulli | estimated IBM fit to Norwegian clinical data |
| Infectiousness Household | | 15% | Bernoulli | estimated IBM fit to Norwegian clinical data |
| Infectiousness Work | | 0.015% | Bernoulli | estimated IBM fit to Norwegian clinical data |
| Infectiousness Nursing Home | | 20.0% | Bernoulli | estimated IBM fit to Norwegian clinical data |
| Infectiousness Generic Contact | | 1.15% | Bernoulli | estimated IBM fit to Norwegian clinical data |
| Maximum daily contacts, mean of normal distribution | | 10 | | estimated IBM fit to Norwegian clinical data |
| Maximum daily contacts, variance of normal distribution | | 3 | | estimated IBM fit to Norwegian clinical data |
| Maximum daily contacts, exponent of power law | | -0.5 | | estimated IBM fit to Norwegian clinical data |

#### Supplementary Table 2. Demographic data used to generate layered network.

In generating the layered network with high-resolution demographic data for each municipality in Norway, we used the data tables from Statistics Norway and from the Norwegian National School Registry (API: https://data-nsr.udir.no/, contains complete information for all schools).

The following Statistic Norway data tables are all available by accession number from https://www.ssb.no/en

| Accession number | Data table text description |
|---|---|
| 3321 | Employed persons (aged 15-74) per 4th quarter, by municipality of work, municipality of residence, contents and year |
| 4469 | Residents in dwellings for nursing and care purposes, by age (M) 2002 - 2019 |
| 6070 | Private households, by type of household (M) 2005 - 2019 |
| 6079 | Private households and persons in private households, by size of household (per cent) (M) (UD) 2005 - 2019 |
| 6206 | Children 0-17 years, by number of siblings and the child's age 2001 - 2019 |
| 6445 | Employed persons, by place of residence, sex and age (per cent). 4th quarter (M) 2005 - 2019 |
| 8947 | Pupils, apprentices, students and participants in upper secondary education, by sex, age and type of school/institution 2006 - 2019 |
| 9169 | Children in kindergartens, by age, hours of attendance per week and ownership (M) 1999 - 2019 |
| 9220 | Kindergartens, by ownership (M) 1987 - 2019 |
| 9929 | Nursing and care institutions and beds, by ownership (C) 2009 - 2018 |
| 10308 | Establishments, by the enterprises sector and number of employees (M) 2012 - 2020 |
| 11933 | Care institutions - rooms, by region, contents and year |
| 12562 | Selected key figures kindergartens, by region, contents and year |

### Supplementary Table 3. List of 15 demographic variables studied in epidemic data from Santé publique

| Variable name | Spearman |
|---|---|
| % households >4 people | 0.54 |
| Population share aged 0-4 | 0.54 |
| Department longitude | 0.53 |
| Population share aged 60+ | -0.53 |
| Population share aged 20-39 | 0.51 |
| Department latitude | 0.48 |
| % households >3 people | 0.46 |
| Household mean people number | 0.40 |
| Department population density / sq. km | 0.38 |
| Populationshare aged 5-19 | 0.36 |
| Department area / sq. km | -0.32 |
| Department population | 0.28 |
| % households >2 people | 0.19 |
| % households >1 people | 0.13 |
| Population share aged 40-60 | -0.02 |

Source for data:

1. Santé publique: Download Sept. 21, 2020 https://www.data.gouv.fr/fr/datasets/donnees-hospitalieres-relatives-a-lepidemie-de-covid-19/
2. French National Institute of Statistics and Economic Studies - Households according to size in 2017. http://www.alisse2.insee.fr/fr/statistiques/2012714
